# Genome-First Approach to Rare and Common Variant Risk of Thoracic Aortic Aneurysm and Dissection

**DOI:** 10.1101/2025.05.18.25327858

**Authors:** John DePaolo, Diane T. Smelser, Dongchuan Guo, Sarah Abramowitz, Gina Sisti, Renae Judy, Nimesh Desai, Wilson Y. Szeto, Michael G. Levin, Hannah Mirshahi, Gregory Salzler, Evan Ryer, James R. Elmore, Scott A. LeMaire, David J. Carey, Dianna M. Milewicz, Scott M. Damrauer

## Abstract

**Background:** Thoracic aortic aneurysm and dissection (TAAD) can have catastrophic health consequences. Eleven genes have strong or definitive evidence for causing heritable TAAD (HTAAD). However, patients are seldom tested for rare pathogenic (P) or likely pathogenic (LP) variants in these HTAAD genes absent a strong family history or syndromic features, and little is known about either their prevalence in the general population or the associated TAAD risk.

Furthermore, the degree to which common genetic variation associated with TAAD modifies rare variant pathogenicity is unknown.

**Methods:** Penn Medicine Biobank (PMBB) and MyCode participants volunteered to have electronic health records linked to biospecimen data including DNA which has undergone genome-wide genotyping and whole exome sequencing. P/LP HTAAD gene variants were adjudicated according to American College of Medical Genetics and Genomics standards, and logistic regression was performed to determine the associated risk of prevalent TAAD. An ascending aortic diameter (AscAoD) polygenic risk score (PRS) was derived from a genome-wide association study (GWAS) of aortic diameter to assess common variant TAAD risk, and regression analyses were performed to determine the modifying effect this PRS had on penetrance of rare variants.

**Results:** Across the two analytic cohorts, 0.2-0.3% of participants carried a P/LP HTAAD gene variant. Compared to individuals without a P/LP variant, carrying a P/LP HTAAD variant was associated with a 13.5-fold increased risk of a diagnosis of TAAD (95% confidence interval [CI] 5.3 to 34.6, *P*<0.001) with variable effects when stratified by gene. A one standard deviation increase in the AscAoD PRS was associated with a 1.43-fold increased risk of TAAD (95% CI 1.39 to 1.47, *P*<0.001). TAAD prevalence was higher among individuals carrying a P/LP HTAAD gene variant in the highest PRS quintile compared to carriers in the lowest PRS quintile (relative risk = 2.32, 95% CI 1.29 to 4.17, *P*<0.01) suggesting that common variant risk may be an important modifier of rare variant risk for TAAD.

**Conclusions:** Our results indicate that P/LP HTAAD gene variants confer a significant increased risk of TAAD in the population at-large, and that polygenic risk may be an important modifier of rare variant risk.

## Introduction

Thoracic aortic aneurysm and dissection (TAAD) comprises both thoracic aortic aneurysm (TAA), which can often go undetected, and thoracic aortic dissection, which occur acutely and are accompanied by catastrophic health consequences. Acute presentations of TAAD have been reported to affect up to 17 per 100,000 individuals per year.^1–6^ Collectively, TAAD cause approximately 8% of out-of-hospital cardiopulmonary arrests annualy.^7^ When detected prior to acute dissection or rupture, elective surgical repair of an aneurysm has superior outcomes with a 30-day mortality rate of 1.7%-3.6% compared to 14-30% for emergency repair.^4,8,9^ Despite understanding the potential dire consequences of undetected or acute cases of TAAD, there are no current screening strategies employed in the population at-large.

Approximately 20-25% of cases of TAAD are heritable.^10^ Historically, TAAD is known to have a Mendelian component; rare pathogenic (P) or likely pathogenic (LP) variants in several known genes associated with heritable TAAD (HTAAD) are associated with familial and/or syndromic disease.^10,11^ Expert consensus has identified more than 30 different genes with varying levels of evidence supporting their causality for HTAAD; 11 of these genes have strong or definitive evidence for causing HTAAD.^10^ Among individuals with sporadic TAAD, 10% have a P/LP variant in an HTAAD gene.^11,12^ However, significant clinical variability has been observed among individuals carrying P/LP variants in HTAAD genes,^13^ and TAAD risk among these individuals in the population at-large has not been adequately characterized.^14^

Increasingly, evidence suggests that common variant genetic risk plays a significant role in cardiovascular disease.^15,16^ Recent genome-wide association studies (GWAS) have identified numerous genetic loci associated with increased ascending and descending thoracic aortic diameter^17,18^ and with TAAD.^19^ To characterize common variant risk, polygenic risk scores (PRS) have been constructed from these GWAS data using different methodologies and have shown consistent association with prevalent and incident thoracic aortic disease in different biobank cohorts. ^17–19^ A PRS for ascending aortic diameter (AscAoD) has been utilized in clinical risk prediction models of aortic dilation, TAA, and dissection in multiple diverse biobanks.^20,21^ Common variant risk is increasingly considered a modifier of monogenic risk in some cardiovascular diseases, especially where significant clinical variability has been observed.^22,23^

In this report, we use whole exome sequencing (WES) linked to electronic health record (EHR) data to investigate the overall risk of carrying a P/LP variant in one of 11 HTAAD genes with strong or definitive evidence for causing TAAD in the Penn Medicine Biobank (PMBB). We then evaluate the contribution of common genetic variation by testing the association of an AscAoD PRS with TAAD events. Finally, we determine the effect of polygenic risk, as proxied by AscAoD PRS, on rare variant risk across the population of individuals carrying a P/LP HTAAD gene variant. Our findings were validated in Geisinger’s MyCode Community Health Initiative (MyCode).

## Methods

### Study Population

The PMBB is a genomic and precision medicine cohort comprising participants who receive care in the Penn Medicine health system and who consent to linkage of electronic health records with biospecimens, including 43,731 with DNA samples that have undergone whole exome sequencing and 43,623 that have undergone genome-wide genotyping (<u>pmbb.med.upenn.edu</u>).^24^ Geisinger is an integrated health care delivery system, and any Geisinger patient of any age can enroll in MyCode. Study participants consent to research linking their biological samples to their medical records. Participants provide blood samples from which DNA is extracted, and sent for whole exome sequencing as previously described in collaboration with RGC.^25^ Similar to previous studies,^19^ in both cohorts TAAD was defined either as ≥2 outpatient or ≥1 inpatient encounters with: 1) the *International Classification of Diseases, 10^th^ Revision* (ICD10) diagnosis code of I71.0, I71.1, I71.2, I71.5, or I71.6, or *International Classification of Diseases, Ninth Revision* (ICD9) codes 441.01, 441.03, 441.1, 441.2, 441.6, or 441.7. EHR data of individuals with confirmed P/LP HTAAD gene variants were reviewed to confirm TAAD diagnoses.

### PMBB Genetic Data

Whole exome sequencing was performed as previously described by Regeneron Genomics Center (RGC).^26^ Individual patient DNA samples were processed and sequenced on the Illumina NovaSeq 6000 (Albany, NY, USA). WeCall variant caller was employed for sequence alignment (GRCh38), variant identification, and genotype assignment.^27^ Quality control exclusions included sex errors, high rates of heterozygosity (D-statistic > 0.4), low sequence coverage, and genetically identified sample duplicates. Single nucleotide variants (SNVs) were filtered for a read depth ≥ 7 and were retained if they either had at least one heterozygous variant genotype with an allele balance ratio ≥ 0.15, or a homozygous variant genotype. Insertion-deletion variants (INDELs) were filtered for a read depth ≥ 10 and either a heterozygous variant genotype with an allele balance ≥ 0.20, or a homozygous variant genotype.

Genotype array analysis was performed as previously described by RGC using a genotyping array chip with 654,027 genetic markers with the addition of ancestry-specific informative markers.^24^ Participant samples were genotyped on Illumina Global Screening Array v.2.0 (GSAv2). Sample level quality control was performed and subsequently imputation was performed using Eagle v2.4.1^28^ and Minimac4 version 1.0.0^29^ software. All autosomes were imputed with TOPMed version R2 on a GRCh38 reference panel. Quality control after imputation included removal of palindromic variants, biallelic variant check, sex check, genotype and sample call rate filtering (>99%), minor allele frequency filtering (MAF > 1%), Hardy-Weinberg equilibrium test (P-value > 1×10^-8^), and imputation score check (R2 > 0.7).^24^ Genetic PCs to adjust for population structure and inform genetically inferred ancestry was performed using EIGENSOFT version 7.2.0.^30^

### MyCode Genetic Data

Sequencing was performed to a read depth of >20x. The Variant Call File sequence reads were aligned to reference genome version GRChr38. Variant quality controls included filtering for quality by depth score >5.0 and combined depth of ≥10 for indels, quality by depth >3 and combined depth of ≥7 for single nucleotide variants, alternate allelic balance >15% (single nucleotide variants) or >20% (indels), and ≥5 alternate reads. Sequence variants were mapped to coding DNA and protein sequence using the National Center for Biotechnology Information RefSeqGene definitions (National Institutes of Health, Bethesda, MD).

Genotype array analysis was performed as part of the GHS-Regeneron Genetics Center DiscovEHR partnership. DNA from participants was genotyped on either the Illumina Infinium OmniExpressExome (8v1-2) or Global Screening Array (GSA) GSA-24v1-0_A1 and imputed to the TOPMed reference panel (stratified by array) using the TOPMed Imputation Server on a GRCh38 reference panel. Before imputation, we retained variants that had a MAF ≥ 0.1%, missingness <1% and Hardy–Weinberg equilibrium test *P* > 10^−15^. After imputation, data from the Infinium OmniExpressExome and GSA datasets were merged for subsequent association analyses, which included an Infinium OmniExpressExome/GSA batch covariate. Genetic PCs were calculated using PLINK (version 2.0).

### Population descriptors

To permit the comparison of genetic effects across diverse populations, individuals were stratified by “population group” corresponding to similarity to 1000 Genomes Project (1000G) reference populations.^31^ To classify individuals to population groups in PMBB, we performed principal component (PC) analysis of participants by projecting participants into 1000G individual PC space based on 1000G continental-level reference populations.^31^ Kernel Density Estimation (KDE) was used to call participant population group based on likelihood similarity. We used KDE to estimate multidimensional PC1 through PC4 for each 1000G continental-level reference population. Participants were then assigned to population groups based on each participant’s highest likelihood relative to the 1000G reference population groups. Analyses were performed among all participants within PMBB, and sensitivity analyses accounted for study-level population stratification by ancestry group.

Population group assignment in MyCode is described in detail in Staples et al.^32^ Briefly, a dataset of the intersecting SNPs from MyCode and the HapMap3 dataset was filtered for common, high quality SNPs (PLINK maf 0.05 --geno 0.1 --snps-only). A KDE was trained for each ancestral superclass from the HapMap3 samples (AFR, AMR, EAS, EUR and SAS), and the likelihood of each sample being in an ancestral subclass was calculated based on the KDEs. MyCode participants were assigned to population superclass based on likelihood.

Borderline samples with 2 or more superclasses were assigned AFR over EUR, AMR over EUR, AMR over EAS, SAS over EUR, AMR over AFR to provide more stringent estimates of EUR, and more inclusive admixed populations.

### HTAAD Variant Calls

Though at least 30 genes have been associated with HTAAD, we selected the 11 genes that have definitive or strong association with HTAAD based on National Institutes of Health Clinical Genome Resource (ClinGen) Aortopathy Working Group, including *ACTA2, COL3A1, FBN1, LOX, MYH11, MYLK, PRKG1, SMAD3, TGFB2, TGFBR1, and TGFBR2*.^10^ Variants identified in these genes were initially filtered for minor allele frequency (MAF) < 0.0001 in the gnomAD database,^33^ and a Rare Exome Variant Ensemble Learner (REVEL)^34^ score ≥ 0.5 for nonsynonymous variants. Subsequently P/LP variants were labeled as pathogenic or likely pathogenic based on American College of Medical Genetics classification framework which relies on observed penetrance of given variants, segregation of a specific variant with cases of disease within a family, and presence of the variant in cases observed in unrelated individuals.^35,36^

### Polygenic risk score creation

We constructed a PRS for AscAoD from a GWAS of AscAoD among EUR UKB participants.^17^ The 1,117,325 variants and their weights were selected using PRS-CS using default settings.^37^ Individual PMBB or MyCode participant scores were calculated using pgsc_calc, a pipeline developed by the PGS Catalog that computes individual scores by combining imputed genotypes with PRS weights.^38^ As allele frequency differences across diverse populations can influence PRS distribution and limit accuracy, pgsc_calc allows a genetic principal component (PC)-based method to normalize scores across populations by adjusting the mean and variance relative to the 1000 Genomes Project and Human Genomes Diversity Project (HGDP).^39^ In this approach, a PC space is created using reference populations and a PRS is modeled as a linear function of the PCs. Score residuals are calculated as the difference between the observed and predicted PRS, and then divided by the standard deviation of the residuals. This ensures that the adjusted PRS values have a mean of 0 when considering the influence of population diversity. To adjust for the variations in PRS range, the variance of the residuals is modeled as a function of the PCs, and then is used to normalize the residual PRS. The variance of the final PRS values distribution is therefore approximately 1 across all populations.

### Statistical Analysis

Firth’s bias-reduced logistic regression was employed to evaluate the association of carrying a P/LP variant in a HTAAD gene with prevalent TAAD, overall and, separately, stratified by genetically similar population group. Where indicated, effects were meta-analyzed across cohorts or populations using inverse variance weighting with fixed effect. Firth’s bias-reduced logistic regression was also used to determine the effect of carrying gene-specific P/LP variants among all participants, and stratified by population group within populations where at least one individual carried a P/LP variant for a specific HTAAD gene. Logistic regression analysis was performed to determine the association of change in AscAoD PRS and TAAD risk. Each regression model was adjusted for age, sex, and the first five genetic PCs. For comparison of effects across cohorts or population groups, heterogeneity between effect estimates was evaluated using χ^2^, I^2^, and τ^2^ tests. The difference in relative risk of TAAD among P/LP variant carriers in the highest versus lowest quintile of polygenic risk was assessed by calculating relative risk using the Mantel-Haenszel test. The marginal effect across the spectrum of AscAoD PRS values, stratified by the presence or absence of P/LP variants in HTAAD genes, was evaluated using the marginaleffects() package version 0.7.0 and visualized with 95% confidence intervals using the plot_predictions function. All statistical analyses were performed using R version 4.2.3 (R Foundation for Statistical Computing, Vienna, Austria).

## Results

### Study population and analysis plan

There were 43,252 participants in the PMBB analytic cohort (**Table 1**) and 123 carried a P/LP HTAAD gene variant. Of these 123 participants with a P/LP HTAAD variant, 13 individuals (11%) were genetically similar to the 1000G African (AFR) reference population, 53 (43%) were female, and the median age at analysis of individuals was 55 years-old (interquartile range [IQR] 38 to 65 years). The number of P/LP variants by HTAAD gene ranged from 0 *PRKG1* carriers to 49 *FBN1* carriers. Among the cohort that did not carry a P/LP HTAAD gene variant, 11,098 individuals (26%) were genetically similar to the 1000G AFR reference population, 21,612 (50%) were female, and the median age at analysis was 57 years-old (IQR 43 to 67).

**Table 1:** Clinical characteristics of individuals in the Penn Medicine Biobank with and without pathogenic/likely pathogenic Heritable Thoracic Aortic Aneurysm and Dissection gene variants.

| PMBB Clinical Characteristics of Individuals with and without a P/LP HTAAD Gene Variant |  |  |  |
| --- | --- | --- | --- |
| Characteristic |  | HTAAD Positive Dataset (N = 123) | HTAAD negative dataset (N = 43,129) |
| Sex, N (%) | Female | 53 (43%) | 21,612 (50%) |
|  | Male | 70 (57%) | 21,517 (50%) |
| Genetically Similar Population Group, N (%) | African (AFR) | 13 (11%) | 11,098 (26%) |
|  | European (EUR) | 105 (85%) | 29,845 (69%) |
| Median Age, years (IQR) |  | 55 (38-65) | 57 (43-67) |
| Median BMI (IQR) |  | 27 (23-31) | 28 (24-33) |
| Median Height in meters (IQR) |  | 1.76 (1.67-1.83) | 1.70 (1.63-1.78) |
| Diagnosis of Hypertension, N (%) |  | 76 (62%) | 24,138 (56%) |
| Diagnosis of Hyperlipidemia, N (%) |  | 46 (37%) | 15,189 (35%) |

Individuals heterozygous for a P/LP HTAAD gene variant were slightly taller and had increased prevalence of hypertension.

There were 163,224 participants in the MyCode cohort (**Supplemental Table 1**). Of the 328 individuals carrying a P/LP HTAAD variant, 13 (4.0%) were genetically similar to the 1000G AFR reference population, 195 (59%) were female, and the median age was 59 years-old (IQR 45 to 73). Among the cohort that did not carry a P/LP HTAAD gene variant, 4,902 individuals (3%) were genetically similar to the 1000G AFR reference population, 101,320 (61%) were female, and the median age at analysis was 63 years-old (IQR 50 to 76).

An overview of the analyses is presented in **Figure 1**. We first estimated the effect of P/LP HTAAD gene variants on risk of prevalent TAAD in both the PMBB and MyCode cohorts. This analysis was performed grouping all P/LP variants together and also by individual gene. Next, we evaluated the role of common genetic variants using a PRS for AscAoD. Finally, we tested the additive effect of rare P/LP variant risk and polygenic risk to evaluate the role of common variant genetic risk as a modifier of monogenic risk.

**Figure 1:**
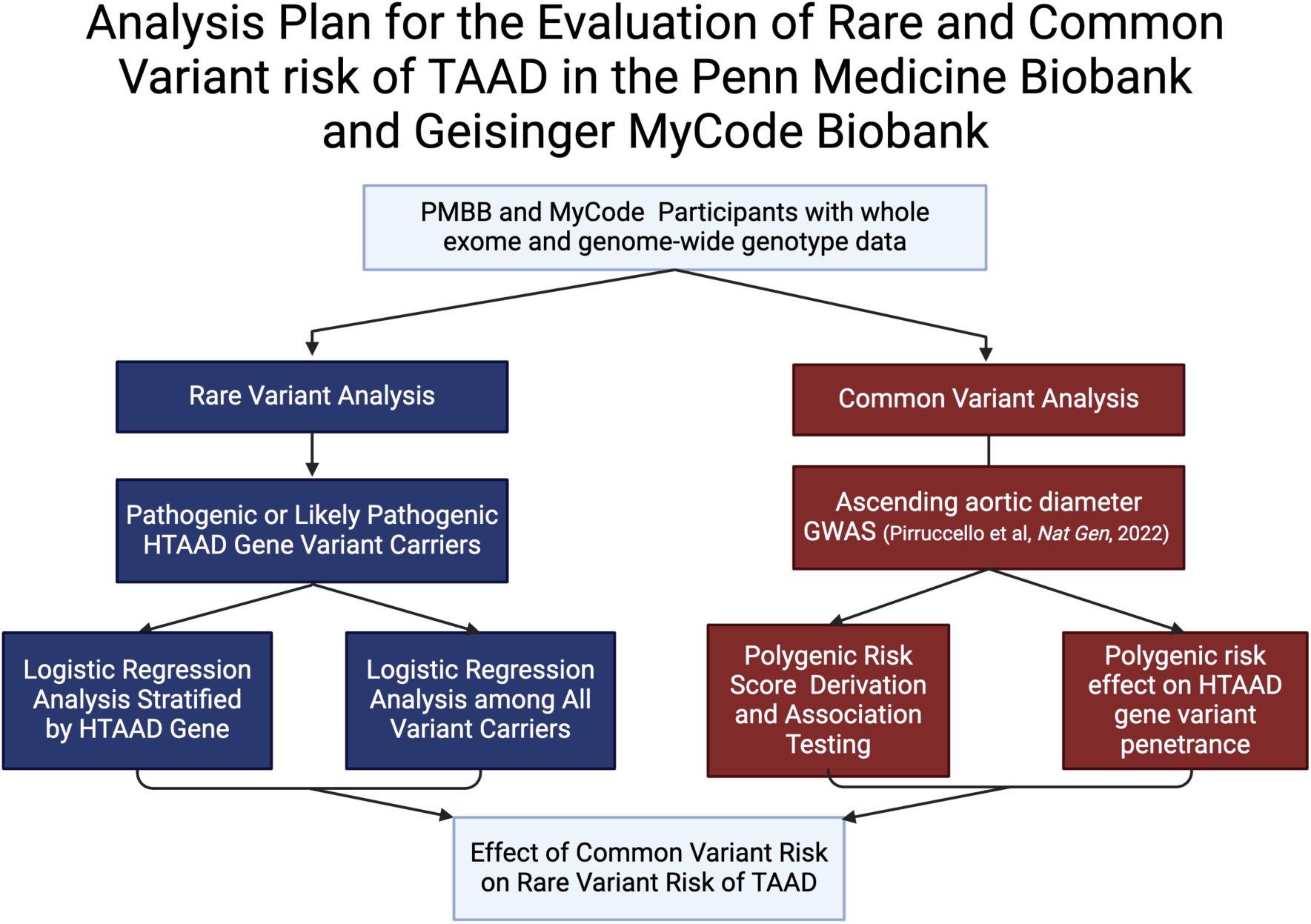
Analytic approach to the evaluation of rare and common variant Thoracic Aortic Aneurysm and Dissection risk in the Penn Medicine Biobank and Geisinger MyCode Biobank.

### Effect of carrying a P/LP HTAAD gene variant

In the PMBB cohort, 52 of 123 (42%) individuals carrying a P/LP HTAAD variant had clinically diagnosed TAAD translating to an odds ratio (OR) of 22.0 (95% confidence interval [CI] 15.0 to 32.1, *P*<0.001). In the MyCode cohort, 51 of 328 (16%) individuals carrying a P/LP HTAAD variant had clinically diagnosed TAAD translating to an OR of 8.4 (95% CI 6.2 to 11.3, *P*<0.001). When meta-analyzed across both cohorts, carrying a P/LP HTAAD variant was associated with a 13.5-fold increased risk of a diagnosis of TAAD (95% CI 5.3 to 34.6, *P*<0.001) [**Figure 2**]. In the more diverse PMBB cohort, individuals genetically similar to the 1000G EUR reference population had a 20.2-fold increased risk of TAAD (95% CI 13.4 to 30.4, *P*<0.001) and individuals genetically similar to the 1000G AFR reference population had an 18.4-fold increased risk of TAAD (95% CI 5.6 to 60.8, *P*<0.001) without heterogeneity across population groups [**Supplemental Figure 1**]. Stratification by population was not repeated in MyCode due to its being overwhelmingly comprised of individuals genetically similar to the 1000G EUR reference populations.

**Figure 2:**
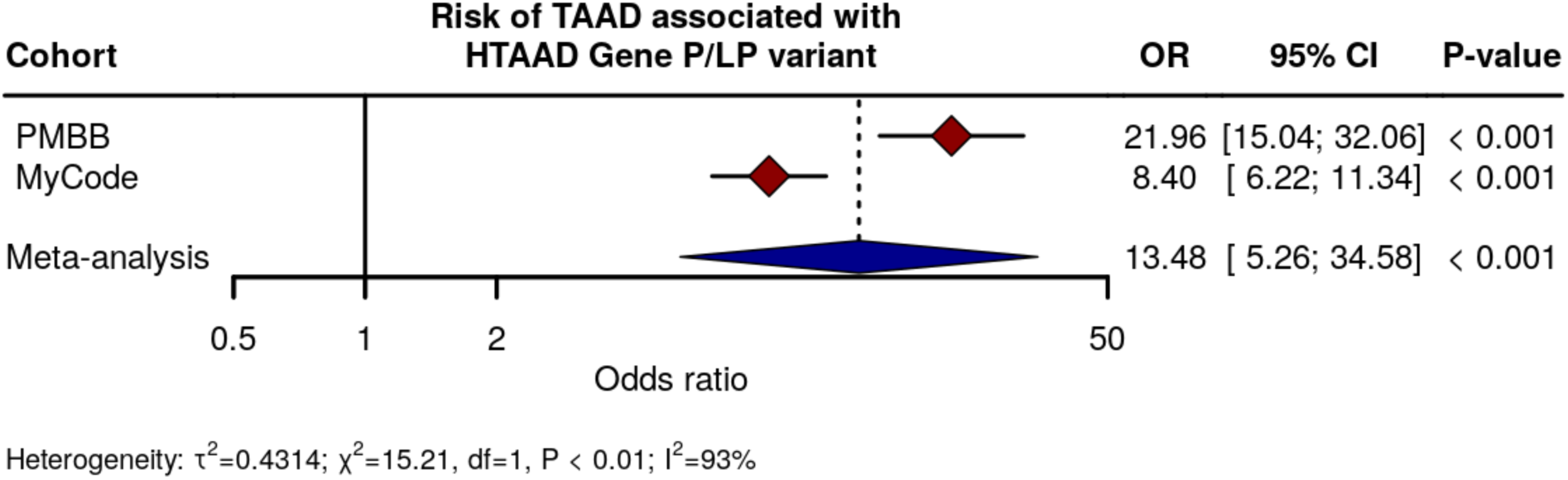
Effect of carrying a rare pathogenic/likely pathogenic HTAAD gene variant on prevalent Thoracic Aortic Aneurysm and Dissection in the Penn Medicine Biobank and MyCode Biobank. Multivariable logistic regression analysis of the association between carrying a P/LP HTAAD gene variant and TAAD adjusting for age, sex, and the first five principal components. CI = confidence interval; HTAAD = heritable thoracic aortic aneurysm and dissection; LP = likely pathogenic; OR = odds ratio; P = pathogenic.

HTAAD gene stratified analyses demonstrated that carrying a P/LP variant in each of HTAAD genes had a statistically significant association with prevalent TAAD (**Figure 3**, **Supplemental Figures 2-11**). *FBN1* was the most common HTAAD gene identified and carrying a *FBN1* P/LP variant was associated with a 73.0-fold increased risk of prevalent TAAD (95% CI 40.0 to 133.3, *P*<0.001). The effect estimates for variants in the other genes ranged from a 6.2-fold increased risk of prevalent TAAD when carrying a *COL3A1* P/LP variant (95% CI 1.5 to 25.6, *P*=0.01) to a 121.4-fold increased risk of prevalent TAAD when carrying a *TGFBR2* P/LP variant (95% CI 4.9 to 2990.6, *P*=0.003). In the MyCode cohort, more individuals (102) carried a P/LP variant in *FBN1* than any other HTAAD gene. The gene-specific effect estimates ranged from 1.4-fold increased risk of TAAD (95% CI 0.3 to 6.9, *P*=0.68) for *COL3A1* to 20.0-fold increased risk of TAAD (95% CI 5.5 to 73.2, *P*<0.001) for *SMAD3* (**Figure 3B**). Taken together, these results demonstrate that carrying a P/LP HTAAD gene variant was associated with a 13.5-fold increased risk of TAAD across the two analytic cohorts, and that the individual gene effects varied substantially.

**Figure 3:**
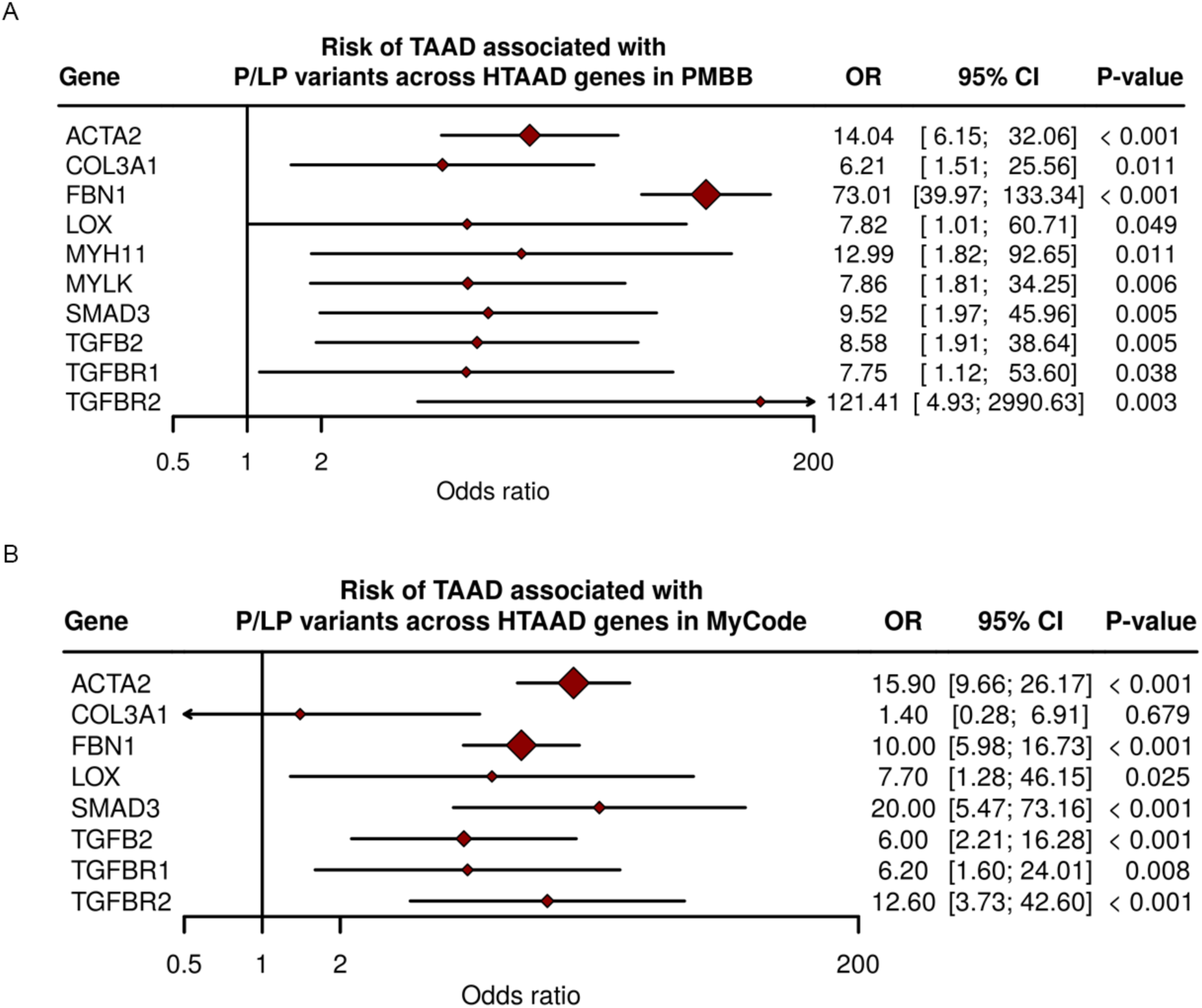
Effect of carrying a rare pathogenic/likely pathogenic HTAAD gene variant on prevalent Thoracic Aortic Aneurysm and Dissection in the PMBB and MyCode biobanks stratified by HTAAD gene. Multivariable logistic regression analysis of the association between carrying a P/LP HTAAD gene variant and TAAD stratified by HTAAD gene. CI = confidence interval; HTAAD = heritable thoracic aortic aneurysm and dissection; LP = likely pathogenic; OR = odds ratio; P = pathogenic.

### Association of polygenic risk with prevalent TAAD

To determine the effect polygenic risk on monogenic P/LP HTAAD gene variant risk, we first applied a PRS for AscAoD to all individuals in the PMBB and MyCode. Individuals with prevalent TAAD had a higher median AscAoD PRS compared to individuals without TAAD (median difference 0.32, *P*=1.5×10^-37^ in PMBB; median difference 0.37, *P*=9.6×10^-85^ in MyCode; **Supplemental Figure 12**). There was a statistically significant association between TAAD prevalence and AscAoD PRS decile in both cohorts (**Supplemental Figure 13**). A one standard deviation (sd) increase in AscAoD PRS was associated with a 1.43-fold increased risk of prevalent TAAD in the PMBB cohort (95% CI 1.36 to 1.50 *P*<0.001) and a 1.44-fold increased risk of prevalent TAAD in the MyCode cohort (95% CI 1.39 to 1.49 *P*<0.001). [**Supplemental Figure 14**]. The meta-analyzed effect across both cohorts was 1.43-fold (95% CI 1.39 to 1.47 *P*<0.001). These results suggest that the AscAoD PRS is associated with increased risk of prevalent TAAD and adequately proxies common variant TAAD risk.

### Effect of common variant risk on rare variant risk

Rare P/LP variants in HTAAD genes are associated with significantly increased risk of TAAD, but there remains substantial clinical heterogeneity among P/LP variant carriers. As an example, only 42% individuals in the PMBB and 16% individuals in MyCode who carried a P/LP HTAAD gene variant had a diagnosis of TAAD. Based on previous investigations in other cardiovascular diseases,^22,23^ we hypothesized that polygenic risk modifies the effect of P/LP HTAAD gene variants.

To further investigate this finding, we stratified individuals by AscAoD PRS quintile and compared the prevalence of TAAD among P/LP HTAAD gene variant carriers in the top quintile of polygenic risk to those in the bottom quintile. In PMBB, the prevalence of TAAD among P/LP HTAAD gene variant carriers in the highest quintile of polygenic was 58% compared to 24% in the lowest quintile of polygenic risk representing a relative risk of 2.43 (95% CI 1.12 to 5.28, *P*=0.02) (**Figure 4A**). In MyCode, the prevalence of TAAD among carriers in the highest quintile of polygenic risk was 20% compared to 9% in the lowest quintile of polygenic risk representing a relative risk of 2.17 (95% CI 0.88 to 5.35, *P*=0.09). When the relative risk difference between highest and lowest polygenic risk quintile was meta-analyzed, there was a 2.32-fold increased risk of prevalent TAAD in the highest quintile of polygenic risk relative to the lowest quintile across both cohorts (95% CI 1.29 to 4.17, *P*<0.01). In both PMBB and MyCode, the marginal effect of a higher AscAoD PRS was increased among the individuals carrying a P/LP HTAAD variant compared to non-carriers. Among P/LP HTAAD gene variant carriers, there was a substantially higher risk of TAAD associated with a higher AscAoD PRS in PMBB (**Figure 4B**) and MyCode (**Figure 4C**). Taken together, these results provide evidence that polygenic risk is an important modifier of monogenic risk and may partially explain the clinical heterogeneity observed in this patient population.

**Figure 4:**
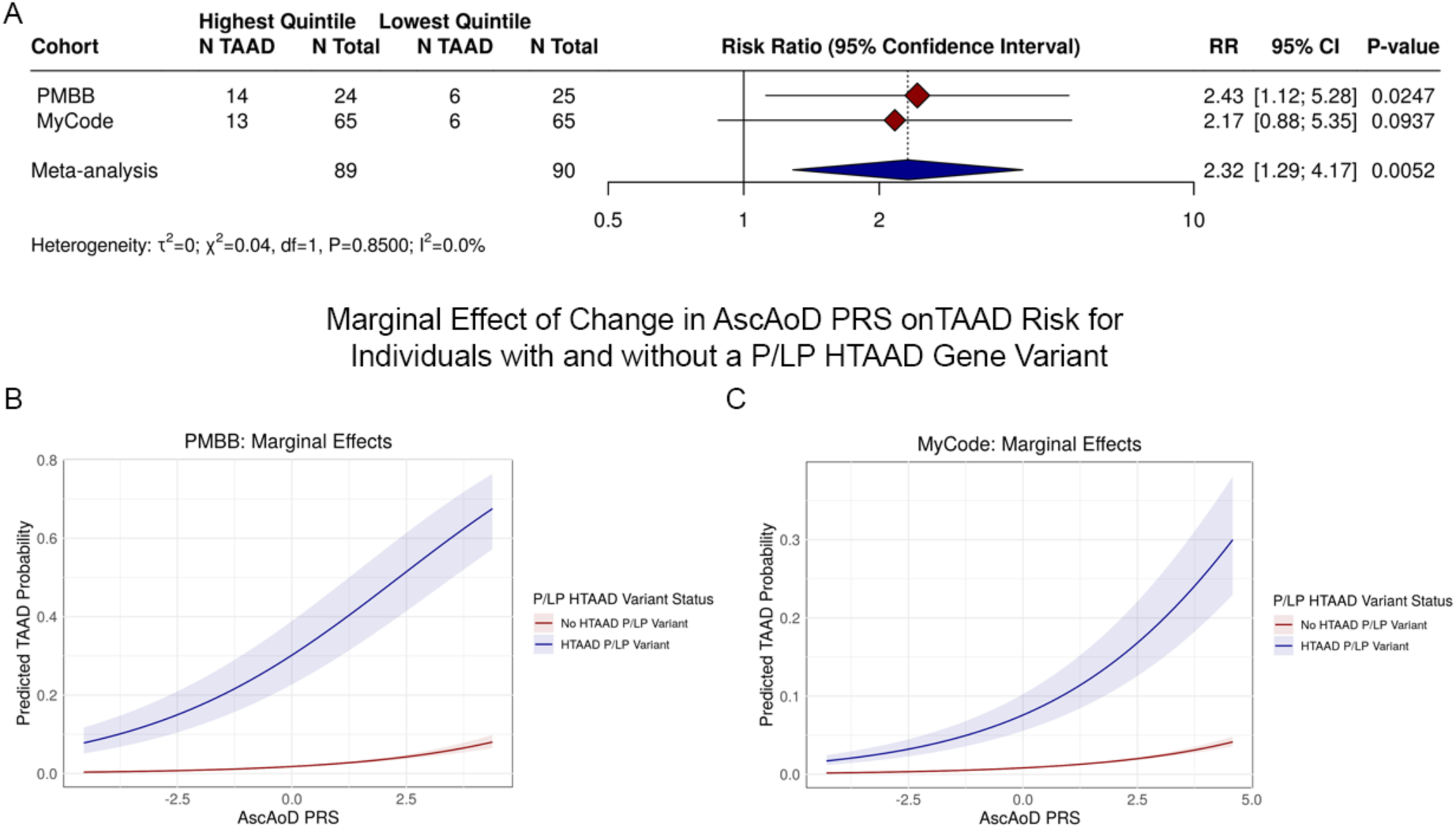
Effect of polygenic risk on rare, monogenic variant risk of thoracic aortic aneurysm and dissection. The AscAoD PRS was applied to individuals in the PMBB and MyCode biobanks who were then stratified into PRS quintiles. **(A)** The prevalence of TAAD diagnoses among individuals carrying a pathogenic/likely pathogenic HTAAD variant in the lowest PRS quintile (N=25 in PMBB, N=65 in MyCode) was compared to TAAD prevalence in the highest quintile (N=24 in PMBB, N=65 in MyCode) to determine the relative risk associated with increased polygenic risk. **(B, C)** Logistic regression analysis was performed to assess the marginal effect of an increased ascending aortic diameter PRS stratified by carrying a P/LP HTAAD variant carrier in either the (B) PMBB or (C) MyCode cohort. The shaded area around both cohorts represent 95% confidence intervals in probability of TAAD. CI = confidence interval; PRS = polygenic risk score; RR = relative risk.

## Discussion

Leveraging data from 2 large biobanks, our results characterize the impact and critical interplay of both rare and common genetic variation on TAAD risk. Our analyses demonstrate that carrying a P/LP variant in one of 11 genes with strong or definitive evidence for causing HTAAD results in an approximately 13.5-fold increased risk of TAAD compared to non-carriers. There was, however, substantial variability in P/LP variant effect across the 11 genes with strong or definitive evidence for causing HTAAD, a result that may have immediate implications for individual-level risk assessment. Our findings also provide evidence for a separate and independent contribution of common genetic, polygenic risk for TAAD and demonstrate the additive nature of these contributions. These results have important implications for estimating and communicating risk associated with HTAAD to patients and their families.

The 11 genes analyzed in this study are included among the ACMG’s list of actionable/returnable variant results because of the strong or definitive evidence of association with HTAAD.^11,40^ However, while previous family-based studies of these genes have signaled extremely high lifetime risk of thoracic aortic disease, even approaching 100% for some *FBN1* variants,^41^ their effect in the general population has been less well characterized. Using biobank data, we were able to estimate the overall risk of TAAD associated with carrying a P/LP variant to be approximately 13.5-fold which provides clinicians with a more generalizable risk estimate, though there is substantial variability in risk across the 11 genes. This is an important advancement in understanding due to the rise of large biobanks and the increasing access to genetic data. These data represent progress in our understanding of the genetic risk for TAAD and will help facilitate more precise counseling for patients and their families. In particular, in an era of emerging genomic medicine these results provide important information for those patients who have incidentally identified HTAAD variants.

A persistent challenge in evaluating individual level risk for TAAD is the variable penetrance,^42^ and several reports have characterized the clinical heterogeneity associated with carrying P/LP variants in HTAAD genes even within the same family.^43–45^ Despite this, there is a limited understanding of the factors influencing disease penetrance. Our work addresses this knowledge gap by demonstrating that common variant risk is a modifying factor in the clinical variability of thoracic aortic disease. In both analytic cohorts we found that there were participants with P/LP HTAAD variants that do and do not carry a diagnosis of TAAD; importantly, those in the highest quintile of polygenic risk had more than twofold increased risk of having TAAD than being in the lowest quintile. Much like environmental risk factors, polygenic risk has an incremental, additive effect on underlying monogenic risk. An individual’s polygenic risk may therefore be an important modifying factor in determining overall TAAD risk related to a P/LP HTAAD gene variant. Our findings provide clinicians a biological explanation for the heterogeneity often observed across the spectrum of individuals with HTAAD.

As thoracic aortic disease is known to have limited warning signs, minimal preventative health or screening strategies, and disastrous health consequences, there have been efforts to develop clinical prediction models for both ascending thoracic aortic dilation and acute aortic events.^46^ Integration of common variant genetic risk into clinical prediction models to determine ascending thoracic aortic dilation risk has yielded modest improvements in their performance among certain patient populations,^20,21^ though there is some evidence that polygenic risk is more effective in modeling dissection.^47^ As GWAS sizes increase, we expect novel common genetic changes to be identified that promote thoracic aortic disease, and the PRSs from these studies to be optimized such that their clinical accuracy and applicability will improve across diverse populations. Eventually, we envision a clinical risk tool that integrates both rare and common variant risk that provides true personalized risk assessment that can guide individual level care for thoracic aortic disease. Such a tool would reshape the current approach to TAAD, optimizing resource utilization while maximizing a precision medicine framework to screening, surveillance imaging, and elective surgical intervention to prevent life-threatening thoracic aortic events.

### Limitations

This work has several limitations. First, although we manually adjudicated outcomes in all 123 individuals with a P/LP HTAAD gene variant in PMBB, we relied upon algorithmically labeled TAAD cases and non-cases in the remainder of the analytic cohorts where there could have been phenotype misclassification that may have introduced bias. Second, while the fidelity of WES has improved dramatically over time, we did not confirm the P/LP variants with Sanger sequencing. Third, though our PRS associates with prevalent TAAD across diverse populations, there is substantial evidence that a PRS constructed from a single population group (in this case the EUR population from UKBB) has poor portability across population groups and may inadequately characterize polygenic risk in non-EUR populations. Future investigations would benefit from using a PRS from a larger, more diverse study of thoracic aortic disease. Finally, while we demonstrated a difference in relative risk of TAAD between the highest and lowest polygenic risk quintiles, we were generally underpowered to appreciate any interaction or synergism between monogenic and polygenic risk. Future investigations of polygenic modification of monogenic risk would benefit from larger cohort studies and from prospective data analysis.

### Conclusions

Carrying a P/LP variant in one of 11 HTAAD genes that have strong or definitive evidence for causing TAAD is associated with a 13.5-fold increased risk of prevalent TAAD. However, the effect of P/LP variants varies substantially across these 11 HTAAD genes.

Polygenic risk is an important modifier of monogenic risk and may have important clinical consequences when determining individual level TAAD risk.

## Abbreviations

AscAoD: Ascending Aortic Diameter
CI: Confidence interval
EHR: Electronic Health Record
HTAAD: Heritable Thoracic Aortic Aneurysm and Dissection
LP: Likely Pathogenic
OR: Odds ratio
P: Pathogenic
PRS: Polygenic Risk Score
RR: Relative risk
TAA: Thoracic Aortic Aneurysm
TAAD: Thoracic Aortic Aneurysm and Dissection
WES: Whole Exome Sequencing

## Data Availability

All data produced in the present study are available upon reasonable request to the authors.

## Acknowledgements

We acknowledge the Penn Medicine BioBank (PMBB) for providing data and thank the patient-participants of Penn Medicine who consented to participate in this research program. We would also like to thank the Penn Medicine BioBank team and Regeneron Genetics Center for providing genetic variant data for analysis. The PMBB is approved under IRB protocol# 813913 and supported by Perelman School of Medicine at University of Pennsylvania, a gift from the Smilow family, and the National Center for Advancing Translational Sciences of the National Institutes of Health under CTSA award number UL1TR001878.

The authors thank all the participants of the MyCode Community Health Initiative Study (MyCode) and the MyCode Research Team as well as the members of the Geisinger-Regeneron DiscovEHR Collaboration. All participants provided informed consent the MyCode Community Health Initiative as approved by the Geisinger Institutional Review Board (IRB # 2006-0258). The aortopathy project described in this article was reviewed and approved by the Geisinger Institutional Review Board.

