## Supplemental Table 1 and Supplemental Figures 1-14 for "Genome-First Approach to Rare and Common Variant Risk of Thoracic Aortic Aneurysm and Dissection"

**Affiliations:**
[1] Department of Surgery, Perelman School of Medicine, University of Pennsylvania, Philadelphia, PA, USA.
[2] Department of Genomic Health, Weis Center for Research, Geisinger Health System, Danville, PA, USA.
[3] Division of Medical Genetics, Department of Internal Medicine, McGovern Medical School, University of Texas Health Science Center at Houston, Houston, TX, USA.
[4] Division of Vascular Surgery, Department of Surgery, Perelman School of Medicine, University of Pennsylvania, Philadelphia, PA 19104, USA.
[5] Division of Cardiovascular Surgery, Department of Surgery, Perelman School of Medicine, University of Pennsylvania, Philadelphia, PA 19104, USA.
[6] Department of Medicine, Division of Cardiology, Perelman School of Medicine, University of Pennsylvania, PA 19104, USA.
[7] Corporal Michael J. Crescenz VA Medical Center, Philadelphia, PA 19104, USA.
[8]Department of Vascular Surgery, Geisinger, Danville, PA 17822
[9] Research Institute and Heart & Vascular Institute, Geisinger, Danville, Pennsylvania, USA.
[10] Michael E. DeBakey Department of Surgery, Baylor College of Medicine, Houston, Texas, USA
[11] Department of Genetics, Perelman School of Medicine, University of Pennsylvania, Philadelphia, PA 19104, USA.
[12] Cardiovascular Institute, Department of Medicine, Perelman School of Medicine, University of Pennsylvania, PA 19104, USA.

* These authors contributed equally to this work

**Address for correspondence:**
Scott Damrauer, MD
Perelman School of Medicine
Division of Vascular Surgery
Hospital of the University of Pennsylvania
3400 Spruce Street, 14th Floor South Perelman Center
Philadelphia, PA 19104


### Supplemental Tables

**Supplemental Table 1:** **Clinical characteristics of individuals in the Geisinger MyCode Biobank with and without pathogenic/likely pathogenic Heritable Thoracic Aortic Aneurysm and Dissection gene variants.**

**PMBB Clinical Characteristics of Individuals with and without a P/LP HTAAD Gene Variant**

| Characteristic | | HTAAD Positive Dataset (N = 328) | HTAAD negative dataset (N = 165,990) |
|---|---|---|---|
| Sex, N (%) | Female | 195 (59%) | 101,320 (61%) |
| | Male | 133 (41%) | 64,661 (39%) |
| Genetically Similar Population Group, N (%) | African (AFR) | 13 (4.0%) | 4,902 (3.0%) |
| | European (EUR) | 309 (94%) | 156,913 (95%) |
| Median Age, years (IQR) | | 59 (44-74) | 63 (50-76) |
| Median BMI (IQR) | | 30 (25.5-34.5) | 30 (25-35) |

### Supplemental Figures

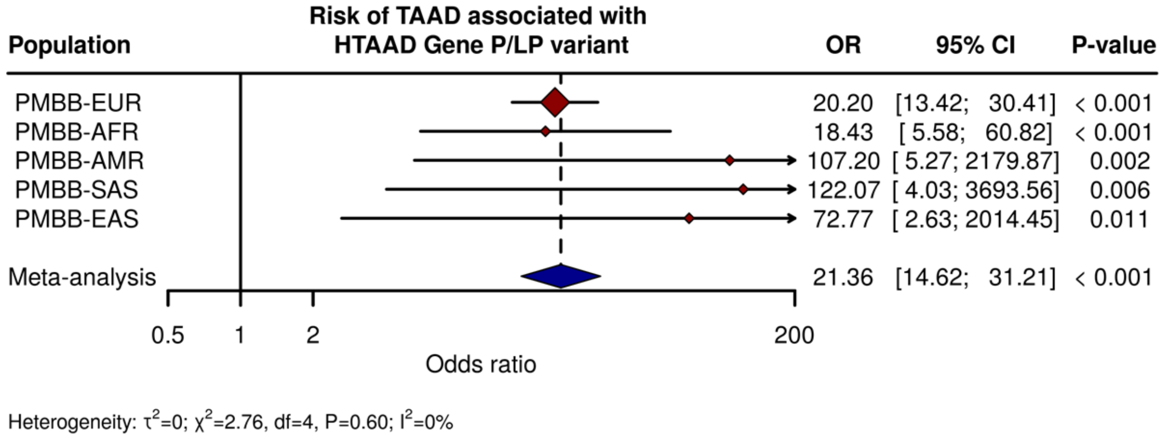


**Supplemental Figure 1: Effect of carrying a rare pathogenic/likely pathogenic HTAAD gene variant on prevalent Thoracic Aortic Aneurysm and Dissection in the Penn Medicine Biobank.** Multivariable logistic regression analysis of the association between carrying a P/LP HTAAD gene variant and TAAD stratified by genetically similar group based on 1000 Genomes Project reference panels. CI = confidence interval; AFR = individuals genetically similar to the 1000G African reference population; AMR = individuals genetically similar to the 1000G Admixed American reference population; EUR = individuals genetically similar to the 1000G European reference population; EAS = individuals genetically similar to the 1000G East Asian reference population; SAS = individuals genetically similar to the 1000G South Asian reference population; HTAAD = heritable thoracic aortic aneurysm and dissection; LP = likely pathogenic; OR = odds ratio; P = pathogenic.

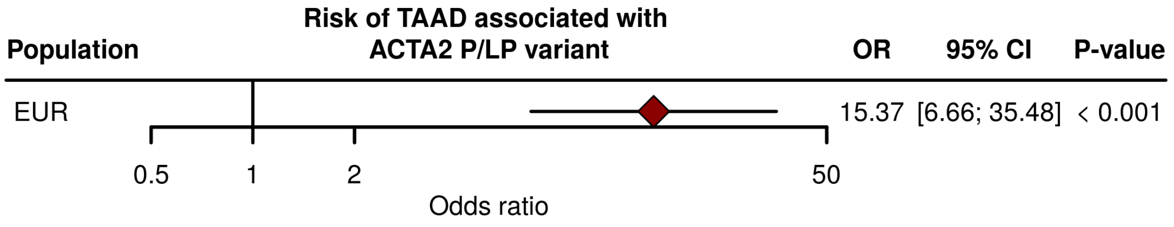


**Supplemental Figure 2: Effect of carrying a rare pathogenic/likely pathogenic *ACTA2* gene variant on prevalent Thoracic Aortic Aneurysm and Dissection in the Penn Medicine Biobank.** Multivariable logistic regression analysis of the association between carrying a P/LP *ACTA2* gene variant and TAAD stratified by genetically similar group based on 1000 Genomes Project reference panels (no individuals genetically similar to the 1000G AFR reference population carried a P/LP *ACTA2* variant). CI = confidence interval; EUR = individuals genetically similar to the 1000G European reference population; HTAAD = heritable thoracic aortic aneurysm and dissection; LP = likely pathogenic; OR = odds ratio; P = pathogenic.

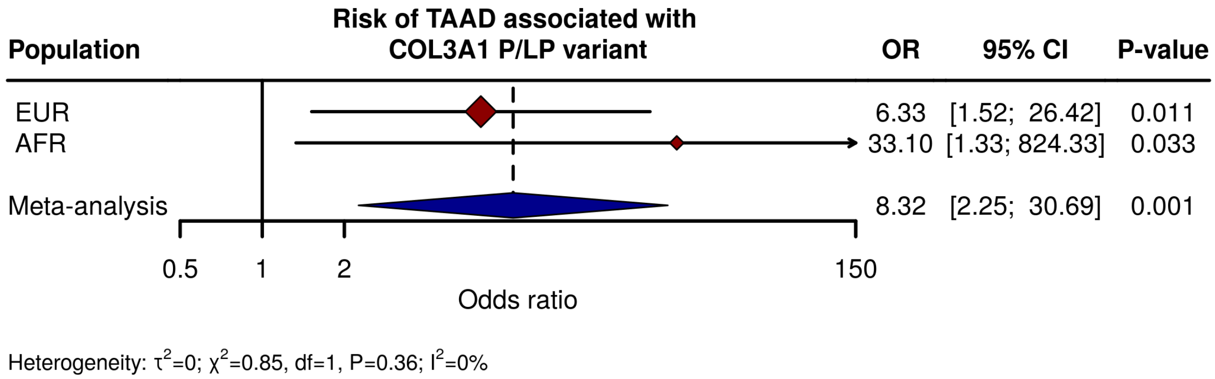


**Supplemental Figure 3: Effect of carrying a rare pathogenic/likely pathogenic *COL3A1* gene variant on prevalent Thoracic Aortic Aneurysm and Dissection in the Penn Medicine Biobank.** Multivariable logistic regression analysis of the association between carrying a P/LP *COL3A1* gene variant and TAAD stratified by genetically similar group based on 1000 Genomes Project reference panels. CI = confidence interval; AFR = individuals genetically similar to the 1000G African reference population; EUR = individuals genetically similar to the 1000G European reference population; HTAAD = heritable thoracic aortic aneurysm and dissection; LP = likely pathogenic; OR = odds ratio; P = pathogenic.

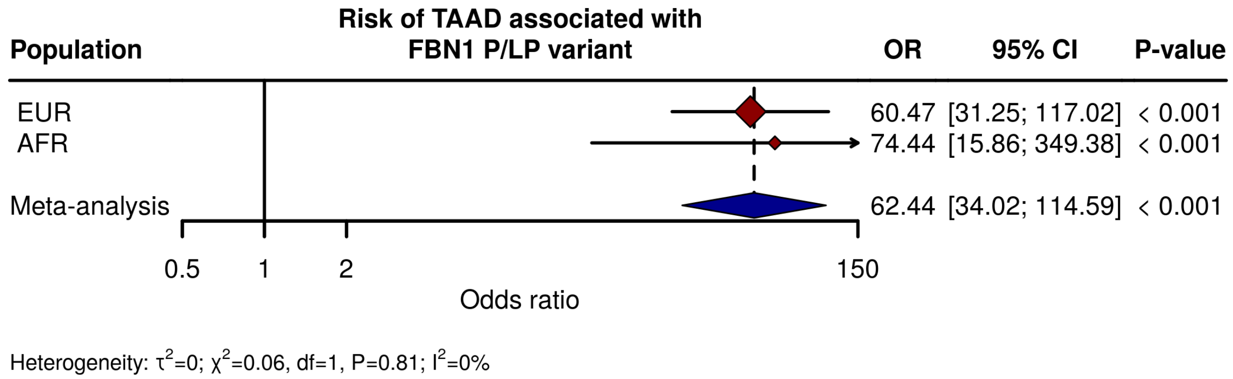


**Supplemental Figure 4: Effect of carrying a rare pathogenic/likely pathogenic *FBN1* gene variant on prevalent Thoracic Aortic Aneurysm and Dissection in the Penn Medicine Biobank.** Multivariable logistic regression analysis of the association between carrying a P/LP *FBN1* gene variant and TAAD stratified by genetically similar group based on 1000 Genomes Project reference panels. CI = confidence interval; AFR = individuals genetically similar to the 1000G African reference population; EUR = individuals genetically similar to the 1000G European reference population; HTAAD = heritable thoracic aortic aneurysm and dissection; LP = likely pathogenic; OR = odds ratio; P = pathogenic.

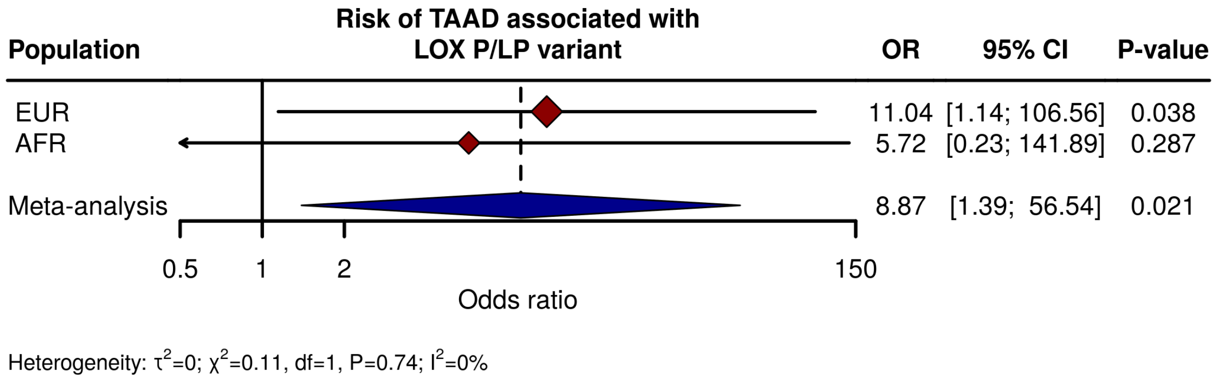


**Supplemental Figure 5: Effect of carrying a rare pathogenic/likely pathogenic *LOX* gene variant on prevalent Thoracic Aortic Aneurysm and Dissection in the Penn Medicine Biobank.** Multivariable logistic regression analysis of the association between carrying a P/LP *LOX* gene variant and TAAD stratified by genetically similar group based on 1000 Genomes Project reference panels. CI = confidence interval; AFR = individuals genetically similar to the 1000G African reference population; EUR = individuals genetically similar to the 1000G European reference population; HTAAD = heritable thoracic aortic aneurysm and dissection; LP = likely pathogenic; OR = odds ratio; P = pathogenic.

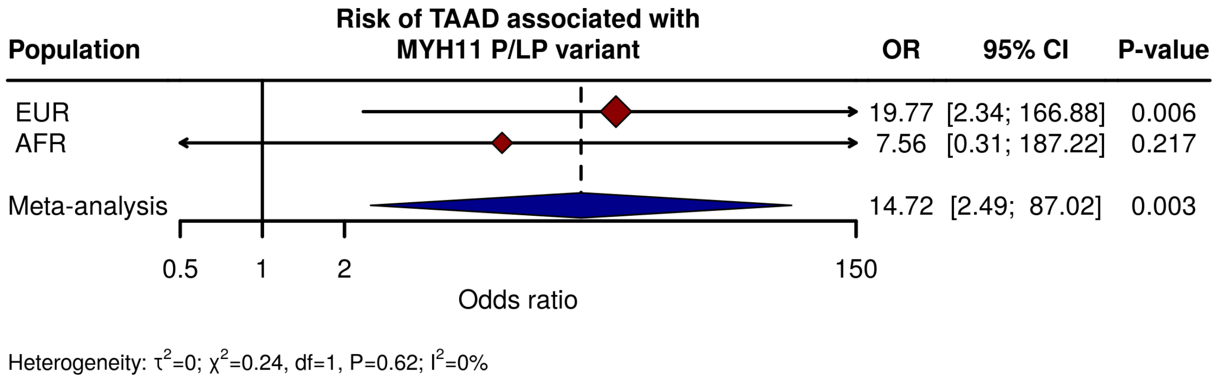


**Supplemental Figure 6: Effect of carrying a rare pathogenic/likely pathogenic *MYH11* gene variant on prevalent Thoracic Aortic Aneurysm and Dissection in the Penn Medicine Biobank.** Multivariable logistic regression analysis of the association between carrying a P/LP *MYH11* gene variant and TAAD stratified by genetically similar group based on 1000 Genomes Project reference panels. CI = confidence interval; AFR = individuals genetically similar to the 1000G African reference population; EUR = individuals genetically similar to the 1000G European reference population; HTAAD = heritable thoracic aortic aneurysm and dissection; LP = likely pathogenic; OR = odds ratio; P = pathogenic.

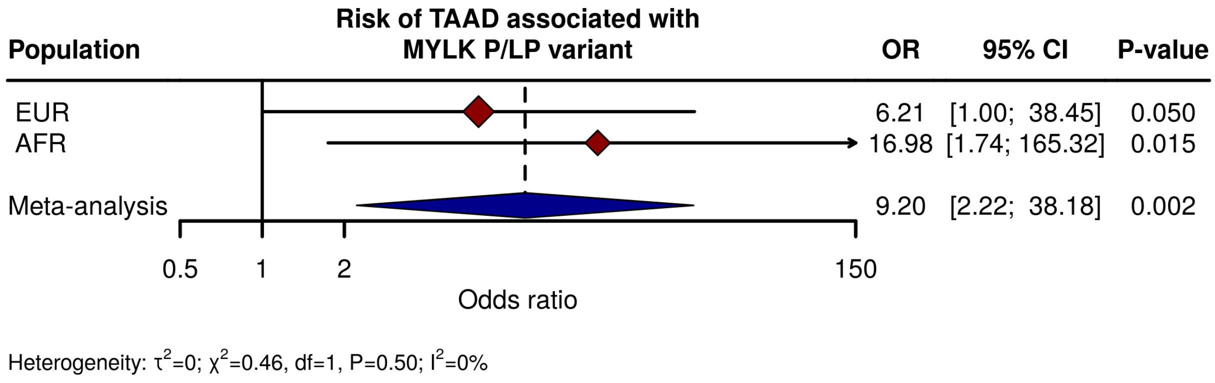


**Supplemental Figure 7: Effect of carrying a rare pathogenic/likely pathogenic *MYLK* gene variant on prevalent Thoracic Aortic Aneurysm and Dissection in the Penn Medicine Biobank.** Multivariable logistic regression analysis of the association between carrying a P/LP *MYLK* gene variant and TAAD stratified by genetically similar group based on 1000 Genomes Project reference panels. CI = confidence interval; AFR = individuals genetically similar to the 1000G African reference population; EUR = individuals genetically similar to the 1000G European reference population; HTAAD = heritable thoracic aortic aneurysm and dissection; LP = likely pathogenic; OR = odds ratio; P = pathogenic.

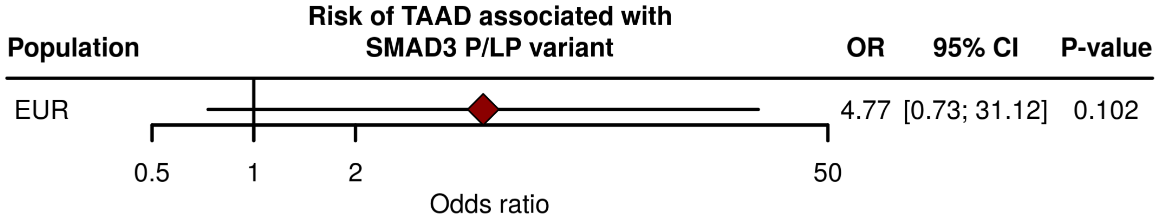


**Supplemental Figure 8: Effect of carrying a rare pathogenic/likely pathogenic *SMAD3* gene variant on prevalent Thoracic Aortic Aneurysm and Dissection in the Penn Medicine Biobank.** Multivariable logistic regression analysis of the association between carrying a P/LP *SMAD3* gene variant and TAAD stratified by genetically similar group based on 1000 Genomes Project reference panels (no individuals genetically similar to the 1000G AFR reference population carried a P/LP *SMAD3* variant). CI = confidence interval; AFR = individuals genetically similar to the 1000G African reference population; EUR = individuals genetically similar to the 1000G European reference population; HTAAD = heritable thoracic aortic aneurysm and dissection; LP = likely pathogenic; OR = odds ratio; P = pathogenic.

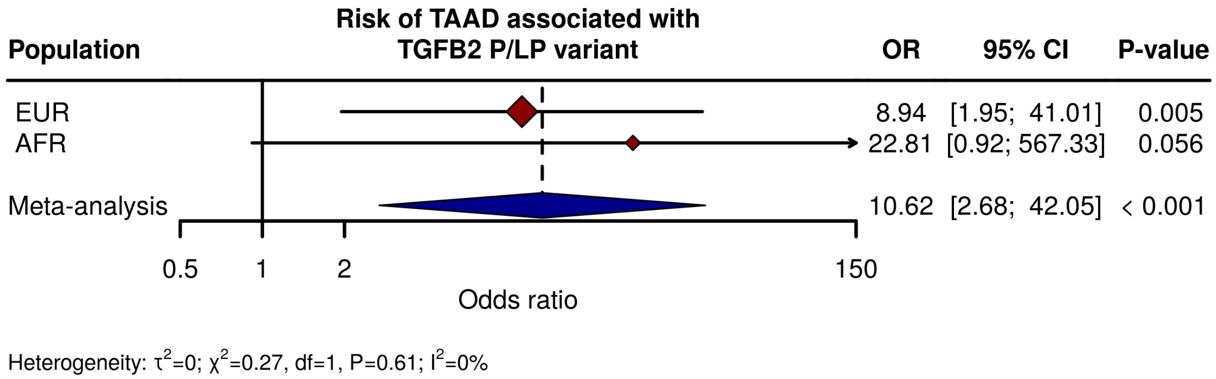


**Supplemental Figure 9: Effect of carrying a rare pathogenic/likely pathogenic *TGFB2* gene variant on prevalent Thoracic Aortic Aneurysm and Dissection in the Penn Medicine Biobank.** Multivariable logistic regression analysis of the association between carrying a P/LP *TGFB2* gene variant and TAAD stratified by genetically similar group based on 1000 Genomes Project reference panels. CI = confidence interval; AFR = individuals genetically similar to the 1000G African reference population; EUR = individuals genetically similar to the 1000G European reference population; HTAAD = heritable thoracic aortic aneurysm and dissection; LP = likely pathogenic; OR = odds ratio; P = pathogenic.

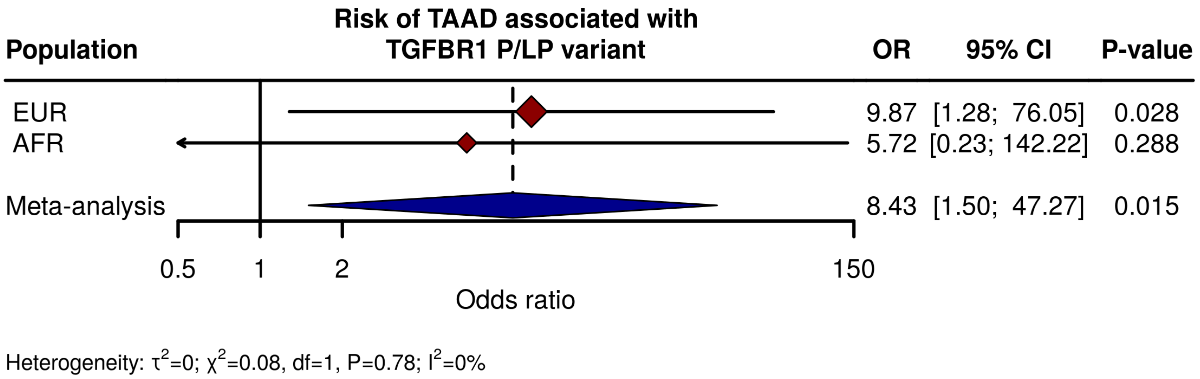


**Supplemental Figure 10: Effect of carrying a rare pathogenic/likely pathogenic *TGFBR1* gene variant on prevalent Thoracic Aortic Aneurysm and Dissection in the Penn Medicine Biobank.** Multivariable logistic regression analysis of the association between carrying a P/LP *TGFBR1* gene variant and TAAD stratified by genetically similar group based on 1000 Genomes Project reference panels. CI = confidence interval; AFR = individuals genetically similar to the 1000G African reference population; EUR = individuals genetically similar to the 1000G European reference population; HTAAD = heritable thoracic aortic aneurysm and dissection; LP = likely pathogenic; OR = odds ratio; P = pathogenic.

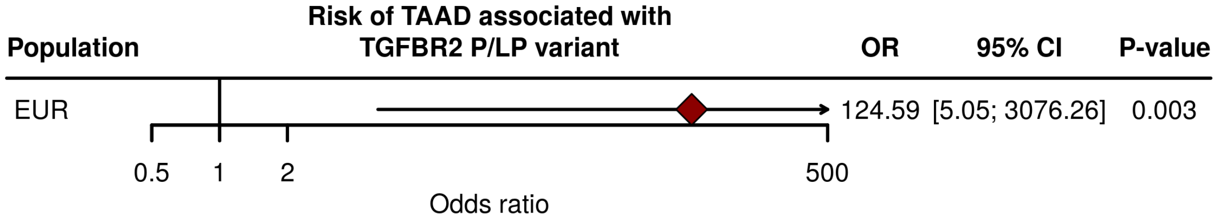


**Supplemental Figure 11: Effect of carrying a rare pathogenic/likely pathogenic *TGFBR2* gene variant on prevalent Thoracic Aortic Aneurysm and Dissection in the Penn Medicine Biobank.** Multivariable logistic regression analysis of the association between carrying a P/LP *TGFBR2* gene variant and TAAD stratified by genetically similar group based on 1000 Genomes Project reference panels (no individuals genetically similar to the 1000G AFR reference population carried a P/LP *TGFBR2* variant). CI = confidence interval; AFR = individuals genetically similar to the 1000G African reference population; EUR = individuals genetically similar to the 1000G European reference population; HTAAD = heritable thoracic aortic aneurysm and dissection; LP = likely pathogenic; OR = odds ratio; P = pathogenic.

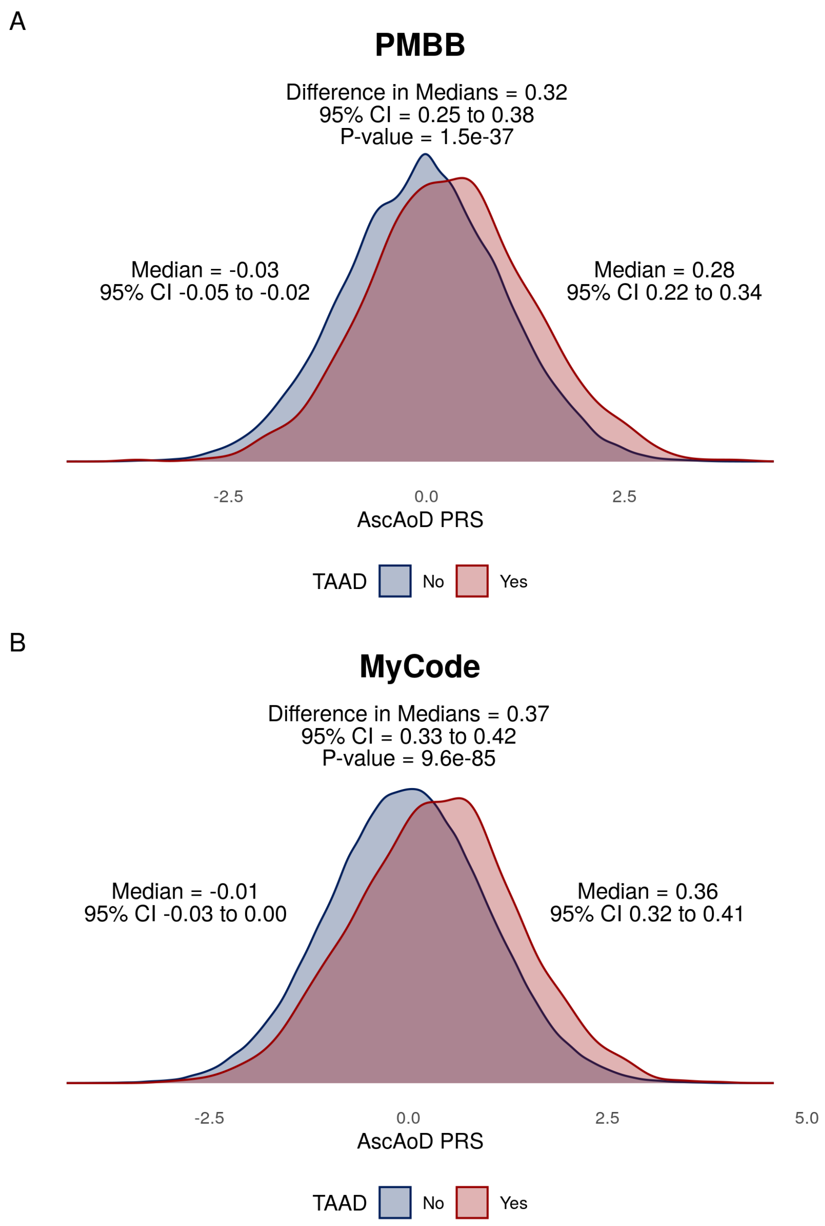


**Supplementary Figure 12: Difference in median ascending aortic diameter PRS between individuals with and without a diagnosis of TAAD.** Density plot of the median AscAoD PRS value of individuals with (red) and without (blue) a diagnosis of TAAD. Difference in median PRS values above the density plots. CI = confidence interval.

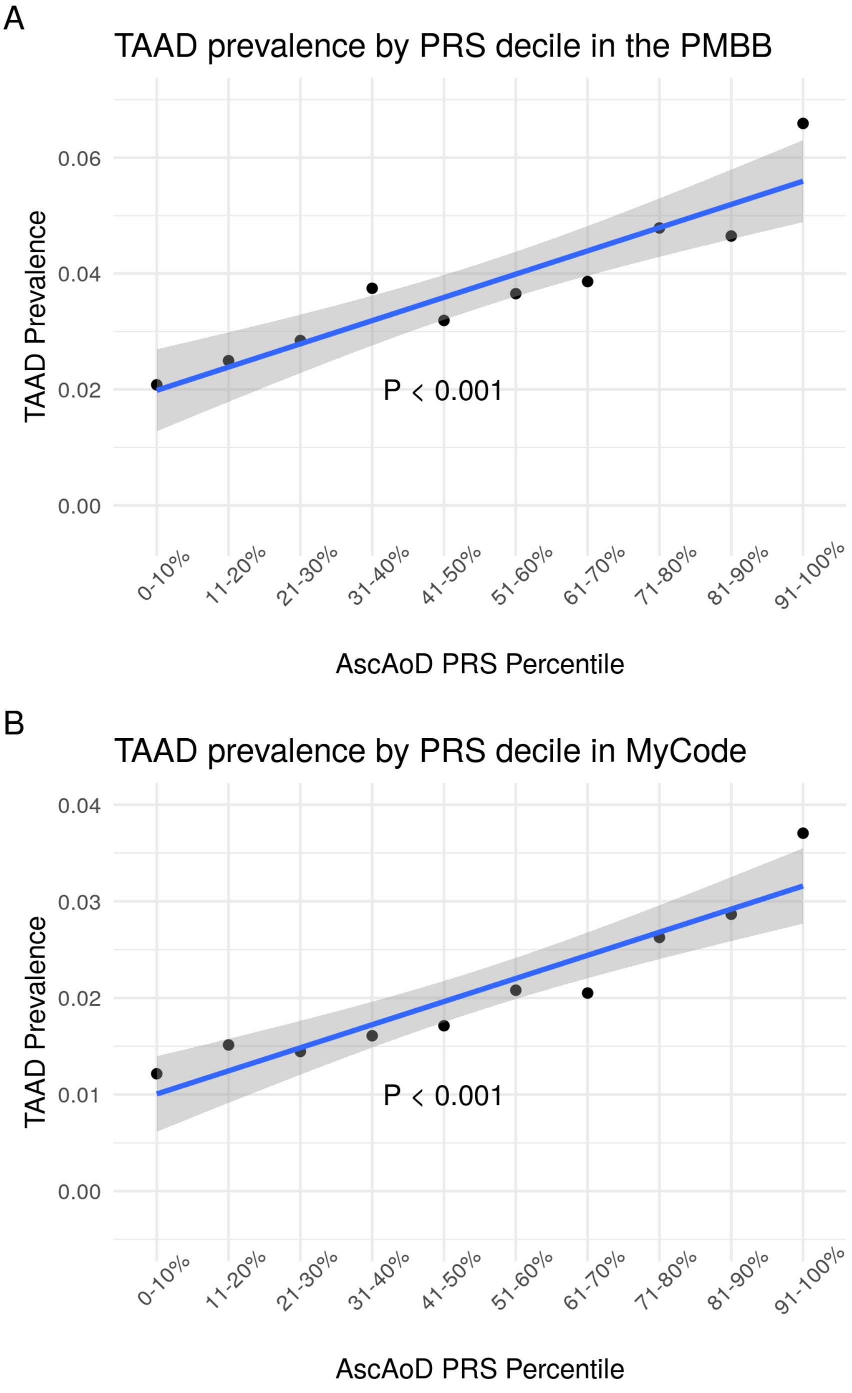


**Supplementary Figure 13: Prevalence of Thoracic Aortic Aneurysm and Dissection diagnoses among Penn Medicine Biobank participants stratified by increasing ten-percentile bins of ascending aortic diameter polygenic risk.**

Ascending aortic diameter PRS was applied to individuals in the Penn Medicine Biobank who were then stratified into ten-percentile groups based on increasing PRS (x-axis) and plotted according to prevalence of TAAD (y-axis) within each group. Each point represents the prevalence of TAAD diagnoses within a specific five-percentile bin, and the blue line represents linear regression analysis of the relationship between PRS percentile bin and TAAD prevalence with error bars equating to the 95% confidence intervals. The grey shaded area corresponds to the 95% confidence intervals of the linear regression.

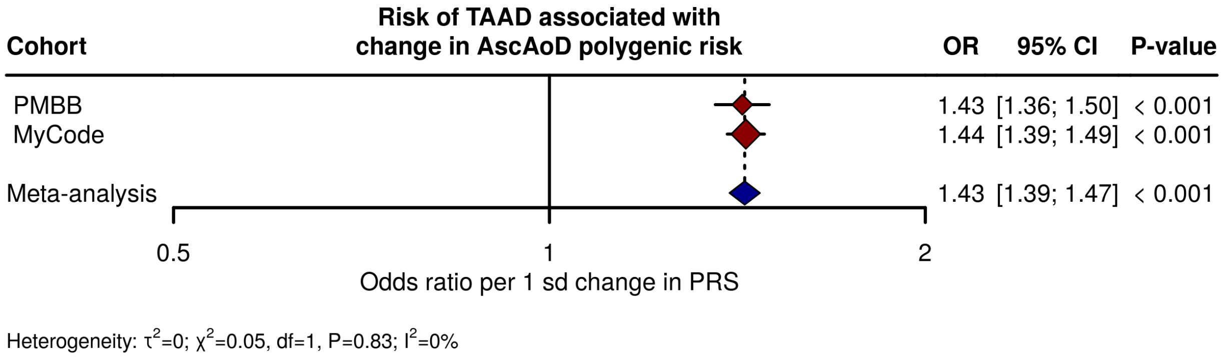


**Supplemental Figure 14: Effect of change in ascending aortic diameter polygenic risk on TAAD risk.** Multivariable logistic regression analysis of the association between one standard deviation change in AscAoD polygenic risk score and prevalent TAAD in the Penn Medicine Biobank. CI = confidence interval; OR = odds ratio; sd = standard deviation.
